# Talking about diseases; developing a model of patient and public-prioritised disease phenotypes

**DOI:** 10.1101/2023.12.19.23300163

**Authors:** Karin Slater, Paul N. Schofield, James Wright, Paul Clift, Anushka Irani, William Bradlow, Furqan Aziz, Georgios V Gkoutos

## Abstract

**Background:** Deep phenotyping describes the use of formal and standardised terminologies to create comprehensive phenotypic descriptions of biomedical phenomena. While most often employed to describe patients, phenotype models may also be developed to characterise diseases. These characterisations facilitate secondary analysis, evidence synthesis, and practitioner awareness, thereby guiding patient care. The vast majority of this knowledge is derived from sources that describe an academic understanding of disease, including academic literature and experimental databases. Previous work has revealed a gulf between the priorities, perspectives, and perceptions held by healthcare researchers and providers and the users of clinical services. A comparison between canonical disease descriptions and phenotype models developed from public discussions of disease offers the prospect of discovery of new phenotypes, patient population stratification, and targeted mitigation of symptoms most damaging to patients quality of life.

**Methods:** Using a dataset representing disease and phenotype co-occurrence in social media text, we employ semantic techniques to identify phenotype associations for a set of common and rare diseases, constituting a phenotype model for those diseases that represents the public perspective. We create an integrated resource for biomedical database and literature-derived disease-phenotype associations by aligning data from several previous studies. We then explore differences between the disease-phenotype associations derived from writing in social media with those from the clinical literature and biomedical databases, with a focus on identification of differential themes and novel phenotypes. We also perform an evaluation of associations for several diseases, with specialist clinicians reviewing associations for validity, feasibility, and involvement in clinical care.

**Findings:** We identified 35,782 significant disease-phenotype associations from social media across 311 diseases, of which 304 could be linked to a combined resource of associations derived from academic sources. Social media-derived disease profiles recapitulated those from academic sources (AUC=0.874 (.95=0.858-0.891)). We further identified 26,081 novel phenotype associations that were not contained in the academic sources, of which 15,084 were considered significant. Constitutional symptoms, those holistic manifestations of disease affecting quality of life, were strongly over-represented in the social media phenotype, contributing more associations especially to endocrine, digestive, and reproductive diseases. An expert clinical review found that social media-derived associations were considered similarly well-established to those derived from literature, and were seen significantly more in patient clinical encounters.

**Interpretation:** The phenotype model recovered from social media presents a significantly different perspective than existing resources derived from biomedical databases and literature, providing a large number of associations novel to the latter dataset. We propose that the integration and interrogation of these public perspectives on disease can inform clinical awareness, improve secondary analysis, and bridge understanding across healthcare stakeholders.

## Background

Deep phenotypes have the power and potential to support unprecedented advances in healthcare [31]. While deep phenotyping is most often employed to construct phenotypic profiles of patients to facilitate personalised and precision medicine, for example in patient population stratification and diagnosis [29, 48], they may also be used to operationally define diseases themselves.

Organised deep phenotypes for diseases were historically developed for databases of rare disease phenotypes manually curated from patient descriptions[22, 52]. With the advent of more informatics-led approaches, these resources were extended to describe common, complex, and rare diseases, with associations between diseases and phenotypes being identified through analysis of data derived from experiments, manual curation, and information extraction from the literature. For example, Hoehndorf et al explored the human diseasome in the context of variant prioritisation and thematic analysis of inter-related disease areas [26], while Kafkas et al linked literature mined phenotype data to BioBank profiles, and another recent literature-based approach was performed by Pilehvar et al, integrating modern natural language processing approaches. These resources provide a rich source of background knowledge, which have proven useful for a wide range of tasks, including differential diagnosis [56, 38] and causative genetic variant prediction [32, 11]. They also provide a data-driven method for explication of diseases, which can be interrogated for supporting clinical awareness of disease symptoms [61], and identifying sub-types in the case of diseases with complex or spectral presentation [3], thereby supporting and facilitating healthcare across multiple stages of translation and implementation.

Nevertheless, the more recent drive for digital phenotyping [31], a sub-field of deep phenotyping that aims to integrate novel data sources, such as wearable devices and social media, into our understanding of disease, has largely not found its way to contributing to those background knowledge resources describing diseases themselves, despite being increasingly popular in patient-level investigations [31]. As such, the background knowledge resources continue to be derived almost exclusively from institutional sources of knowledge, largely academic literature and experimental data. Scientific knowledge resources reflect scientific interest, which is influenced by cultural trends, funding availability, and personal interest [46]. This leads to an imbalance in development of knowledge, with scientific attention not always being fully aligned with need or equity. These scientific knowledge resources form the substrate from which treatment guidelines are synthesised, and thereby from which medicine is practised.

Similar imbalances occur in direct healthcare practice, where social biases play out in medical training and practice, generating disadvantages for sub-populations, for example on the basis of gender, race, age, geography, and economic class. Many such groups have characteristic health problems which are consequently addressed poorly, or not at all. For example, the tendency for medical professionals to ignore or downplay reports of symptoms by women is widely reported and explored in literature [66]. These issues are associated with reduced quality of care, extremely long delays in the diagnosis of diseases, for example endometriosis [1], or causing a large proportion of cardiologists self-reporting as unprepared to diagnose cardiovascular disease in women [8]. In a related example, exposure to topics of transgender health is extremely low in medical curricula [16]; this is then manifest in the experience of those individuals facing high levels of discrimination seeking healthcare [65] and in poor routine data collection negatively affecting resources that are used for medical research [43]. These issues are compounded by the inherent limitations in statistical power when considering minority cohorts [42]. In some cases, these issues translate to more limited understanding of the diseases from which these groups are more likely to suffer [40]. A recent study reports a strong preference amongst clinicians for downgrading the importance of patient-reported symptoms, mis-attributing them either through misunderstanding of terms used or pre-judgement, particularly with constitutional symptoms [58], emphasising the existence of an underlying semantic mismatch between patient and health professional discourse.

The combination of such limitations in scientific exploration and medical care are deeply inter-linked and self-perpetuating. If an incomplete understanding of a disease informs experimental design, clinical care, and clinical data collection, these data and outcomes are then used to inform new academic research, and in turn the clinical practice informed by it. Fundamentally, this feedback loop limits the ability to discover and integrate new perspectives that could lead to improved understanding and treatment of diseases. In addition, previous research has shown that patients and clinicians hold different perspectives and priorities concerning disease [14, 64]. For example, patients usually consider news of a ‘stable’ cancer as being negative, while doctors often consider this to be positive [9]. Meanwhile, analysis of an online Uveitis patient forum identified different language and priorities than those expressed by existing scientific and clinical knowledge resources [50].

To bridge these gaps in understanding of diseases, the patient-centred care paradigm aims to integrate the perspectives and priorities of patients and the public into clinical care [59]. Patient Reported Outcome Measures and Quality of Life assessments are methods used to identify patient perspectives and to reconcile them with clinical understanding. However, patient and public-centred approaches to developing knowledge resources and phenotype models for clinical practice and research have been under-explored. Lenzi et al used a topic modelling approach to discern a digital phenotype for diabetes based on an Italian patient forum. Meanwhile, Maggio et al focused on discussion of methodological aspects of digital phenotyping from social data, and linked data retrieved from social media to epidemiological data. However, existing approaches focus on single diseases, do not correlate their knowledge with literature resources in a systematic way, provide a framework for hypothesis generation for novel associations, and do not provide an open database for secondary research use. Meanwhile, the approaches that focus on creating phenotypes for large numbers of diseases, in addition to being derived entirely from literature and biomedical resources, are rarely actively evaluated in a real clinical context.

In this work, we propose an approach to developing a phenotype model to represent and expose patient and public perspectives and knowledge on disease. We develop a social media phenotype model that identifies disease-phenotype associations for a range of common and rare diseases. We hypothesise that the perspectives presented by this resource will significantly differ from existing knowledge resources. To test this hypothesis, we create a consolidated Biomedical Database and Literature Phenotype with which to compare and contrast the social media-derived associations, highlighting novel associations and differences in theme. We then perform a clinical evaluation of associations for several diseases, including an evaluation of feasibility of novel phenotype associations found in public discourse, but currently unknown to, or underappreciated in the literature and clinical practice.

## Results

We analysed 68,319,325 records of social media posts provided to us by White Swan that describe mentions of a set of 488 diseases, linked to Disease Ontology (DO) [21, 54] classes based on mentions of their labels in social media text. Table 1 shows that the posts were sourced from a range of social media, mostly comprised of Twitter, Reddit, with smaller contributions from online fora and review narrative. A total of 5,895 unique phenotypes, linked to the Human Phenotype Ontology (HPO) [39] were mentioned across all posts. From these data, we developed the Social Media Phenotype (SMP), which describes 52,198 positive associations for 311 diseases, with 620 unique phenotypes appearing across all associations. Of those, we consider 35,381 to be significantly associated, using an acceptable false discovery rate of 0.0005. We also identified a Biomedical Database and Literature Phenotype (BDLP) by combining disease-phenotype associations defined by multiple works from a combination of structured databases and literature-based text analysis. To facilitate a comparative analysis of these phenotype models, we linked profiles for 304 diseases from the SMP to their equivalents in the BDLP.

**Table 1.** Document count and contribution by source

| Source | Document Count | Contribution to Total |
| --- | --- | --- |
| Twitter | 51,621,597 | 76% |
| Reddit | 13,712,022 | 20% |
| Boards (online forums) | 2,607,165 | 4% |
| Reviews | 378,541 | 1% |

We created an online resource for exploring and comparing the SMP and BDLP, which is freely available at https://phenotype.digital/.

### Comparative Analysis of SMP and BDLP

Across the 304 linked diseases, social media phenotype profiles strongly recapitulated the BDLP, with a disease reclassification analysis revealing an AUC of 0.874 (.95=0.858-0.891). The SMP describes 25,447 novel phenotype associations: those not appearing in the disease-matched profile in the BDLP. Of those, 15,084 were considered statistically significant, and our subsequent analysis is limited to this subset (except where explicitly labeled).

The DO and HPO organise diseases and phenotypes into a hierarchical classification structure. For example, lower back pain and back pain can both be considered kinds of pain. This allows us to organise the diseases and phenotypes into groups. Table 2 lists the ten most common groups of phenotypes appearing in novel associations, excluding very general phenotypes. Pain was the most frequently appearing group, with 6% of all novel phenotype associations being in this group.

**Table 2.** Top ten most frequently associated groups of novel phenotypes in the SMP. Proportion of novel phenotypes refers to the proportion of phenotypes in the SMP dataset that were not included in the BDLP. For example, 6% of all novel phenotype associations were either pain or more specific kinds of pain (e.g. abdominal pain). Groups of phenotypes with low information content, below 0.6 Resnik, were excluded.

| Phenotype | Inclusion |
| --- | --- |
| Pain (HP:0012531) | 0.06 |
| Functional abnormality of the gastrointestinal tract (HP:0012719) | 0.05 |
| Abdominal symptom (HP:0011458) | 0.03 |
| Abnormality of reproductive system physiology (HP:0000080) | 0.03 |
| Abnormality of blood circulation (HP:0011028) | 0.03 |
| Internal hemorrhage (HP:0011029) | 0.03 |
| Abnormal cell morphology (HP:0025461) | 0.02 |
| Vascular skin abnormality (HP:0011276) | 0.02 |
| Abnormal emotion/affect behavior (HP:0100851) | 0.02 |
| Abnormal myeloid cell morphology (HP:0020047) | 0.02 |

The hierarchical structure of the ontologies also allows us to explore the associations according to the very general groupings of diseases and phenotypes that sit at the top of these structures, such as respiratory phenotypes or metabolic diseases. Figure 2 shows the representation of phenotype associations according to those categories in the HPO and DO. Figure 2a shows a comparison of associations for SMP and BDLP falling under each category in HPO, with constitutional symptoms, as defined by HPO, being clearly over-represented relative both to other facets in the SMP, and to all facets in the BDLP. Other far smaller differences between the two sets are revealed in a greater expression of voice, thoracic cavity, and blood phenotypes in the BDLP, and a greater proportion of growth anomaly and digestive phenotypes appearing in the SMP. These proportions are reflected also across the novel subset of the SMP, shown in Figure 2b, which shows a largely similar distribution of facet expression to that of the full SMP. Figures 2a and 2b show that the distribution of novel phenotypes in SMP is very similar to the overall distribution of phenotypes in BLP. Figure 2c shows the the proportion of novel phenotypes in the SMP accorded to each disease category defined by the Disease Ontology, projected onto the distribution of the membership of the total 311 diseases, showing that these are strongly correlated, though with a greater focus on digestive diseases, and a lesser focus on infectious and mental health diseases

Figure 1 shows the correlative proportions of novel phenotypes for disease and phenotype categories. Of these, 14 were able to be linked by shared anatomical system, such as ‘nervous system phenotypes’ and ‘nervous system diseases.’ Some of the most strongly represented pairs were matching, such as nervous system, with a log occurrence of 6.72, and neoplasm with 6.52. The heatmap also shows a paucity of novel associations for thoracic cavity and voice across all disease categories, though thoracic diseases show some positive relationship with endocrine and integumental phenotypes. Likewise, novel associations for physical disorders were mostly concentrated in nervous system phenotypes, with some contribution also from head or neck.

**Figure 1.**
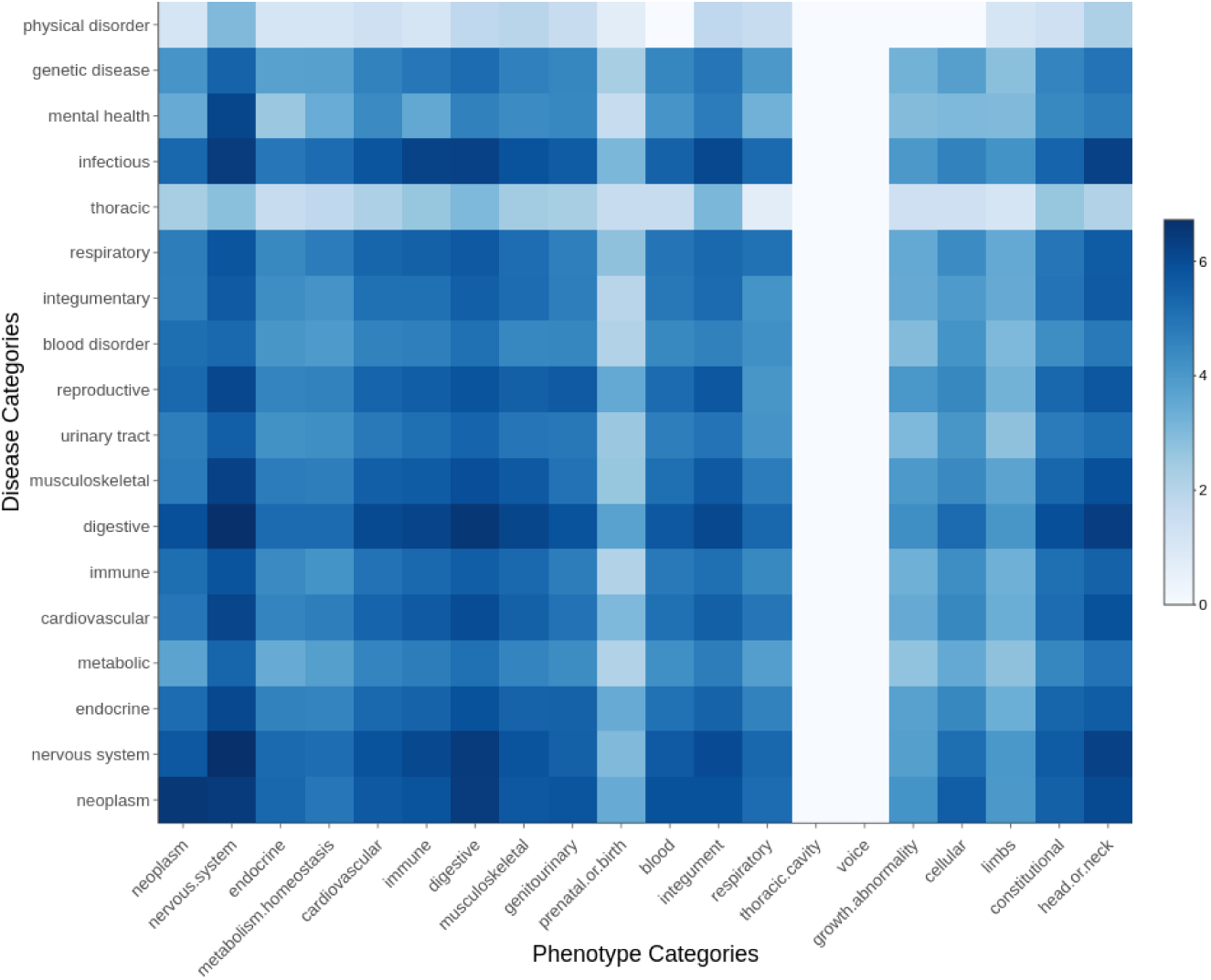
Heatmap displaying the normal log occurrence of novel associations across disease and phenotype categories. The 14 pairs of disease and phenotype categories that refer to matching biological systems are placed symmetrically, and lead the rows and columns.

Due to its clear over-representation in the SMP dataset, we further investigated associations that fell under the constitutional symptoms facet. There are 3,693 constitutional symptom associations in the BDLP, and 3,552 in the SMP, with 2,199 of those being novel (not included in the BDLP). Figure 2d shows the differential distribution of constitutional phenotype associations for the BDLP and SMP across disease categories, proportional to the total number of the linked diseases in each category. This shows that there is a greater focus on constitutional symptoms in the BDLP for thoracic, genetic, cardiovascular, physical, and to a lesser extent nervous system, disorders. Meanwhile, the SMP exhibits a greater focus on constitutional symptoms in reproductive, endocrine, digestive, and integumentary diseases. Across all constitutional symptom associations in both sets, there are only 86 unique classes, with a total of only 28 unique classes appearing across the SMP, with all 86 appearing in the BDLP. Table 3 shows the top groups of associated phenotypes in the BDLP and SMP. In both phenotype models, pain is by far the most compositional constitutional symptom group (BDLP 68% SMP 72%), with other contributions from impairment of activities of daily living, impaired continence, and dyspepsia. The distribution of phenotypes is mostly similar across the two sets of phenotypes, with notable increases in the proportion of back pain (+9%), lower limb pain (+4%), dyspepsia (+4%), and sciatica (+4%) in the SMP. Conversely, the BDLP contains notably greater proportions of chills (+4%). Figure 3 shows a projection of pain phenotype associations for BDLP and SMP, showing clearly that in the SMP, phenotype associations are far more strongly concentrated on a smaller number of phenotypes, with associations being more distributed cross the entire sub-graph in the case of BDLP.

**Figure 2.**
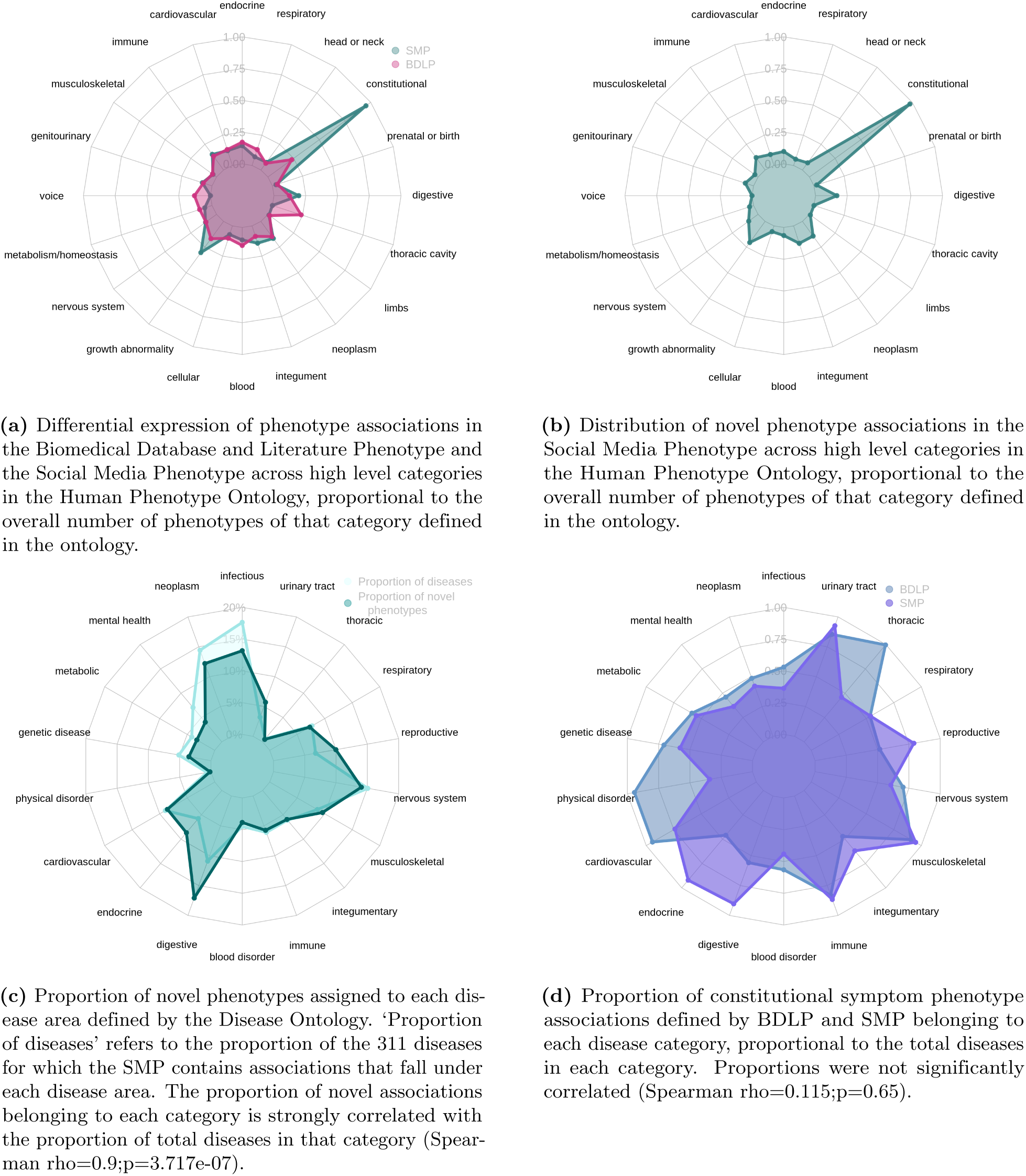
Thematic analyses of phenotype associations in the BDLP and SMP according to high level organisational categories defined by the Human Phenotype Ontology and the Disease Ontology. Phenotype and disease membership in categories is not mutually exclusive: for example, ‘lung cancer’ may be considered both a neoplasm and a respiratory disease.

**Figure 3.**
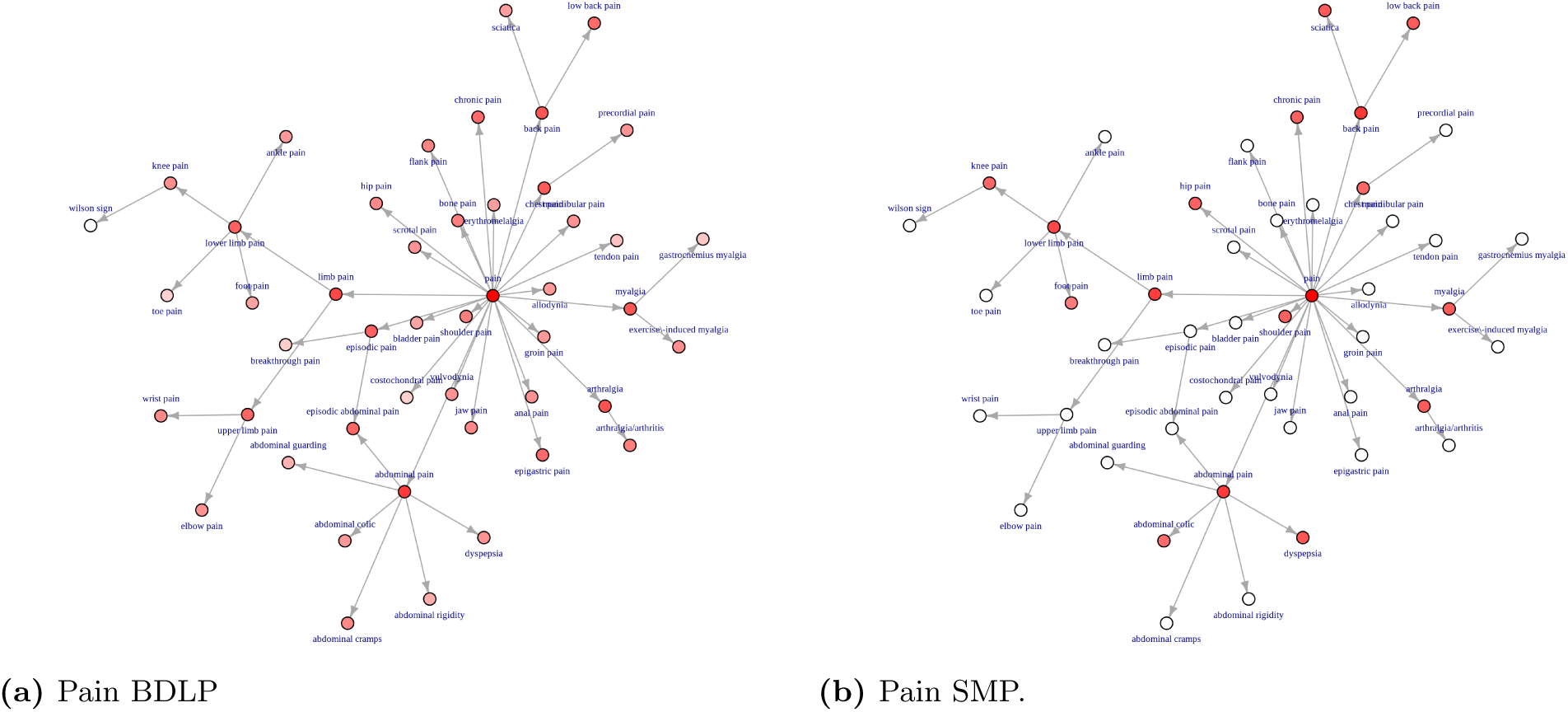
Network projection of pain phenotype associations for the BDLP and SMP. The graph indicates fewer phenotypes represented in the SMP, with stronger focus on more general, or high level, phenotypes that are more closely linked to pain, with associations being more widely distributed across the full set of nodes in the BDLP.

**Table 3.** Top constitutional phenotypes found in the BDLP and SMP. Propertion of associations is the percentage of constitutional symptom phenotype associations that were equivalent to or more specific than the named phenotype. Criteria for inclusion was at least 0.04 proportion of constitutional symptoms in either SMP or BDLP.

| Class | Proportion of associations |  |
| --- | --- | --- |
|  | BDLP | SMP |
| Pain (HP:0012531) | 0.68 | 0.72 |
| Abdominal pain (HP:0002027) | 0.11 | 0.12 |
| Limb pain (HP:0009763) | 0.08 | 0.12 |
| Fatigue (HP:0012378) | 0.07 | 0.05 |
| Impairment of activities of daily living (HP:0031058) | 0.06 | 0.05 |
| Arthralgia (HP:0002829) | 0.05 | 0.05 |
| Back pain (HP:0003418) | 0.05 | 0.14 |
| Myalgia (HP:0003326) | 0.05 | 0.04 |
| Impaired continence (HP:0031064) | 0.05 | 0.05 |
| Chest pain (HP:0100749) | 0.04 | 0.03 |
| Chills (HP:0025143) | 0.04 | 0.0 |
| Lower limb pain (HP:0012514) | 0.04 | 0.08 |
| Pain in head and neck region (HP:0046506) | 0.04 | 0.04 |
| Night sweats (HP:0030166) | 0.02 | 0.05 |
| Dyspepsia (HP:0410281) | 0.01 | 0.05 |
| Urinary incontinence (HP:0000020) | 0.02 | 0.05 |
| Low back pain (HP:0003419) | 0.02 | 0.05 |
| Sciatica (HP:0011868) | 0.01 | 0.05 |
| Neck pain (HP:0030833) | 0.01 | 0.04 |

### Clinical Review and Hypothesis Generation

12 clinical reviewers evaluated phenotype associations for 13 diseases, in total returning 13,088 question responses. The first question evaluated how valid they thought the association was (Figure 4a), with strongly correlated distributions (X^2^ = 47.504 p=1.198e-09), with a slightly greater proportion of ‘not established and unlikely’ results. The second question concerned the type of phenotype association (Figure 4b), and the distribution of responses were also strongly correlated (X^2^ = 76.435, p-value = 4.667e-15). However, this question contains very different response proportions for certain categories. ‘Other associated phenotype,’ was much more common for BDLP associations (0.156) than for those in the SMP (0.052), while ‘Comorbidity,’ and ‘Unknown,’ were seen far more often in response to SMP associations. The third question queried how often the clinicians saw patients with the phenotypes. SMP phenotypes were seen more often in clinic than those in the BDLP (one-tailed Wilcoxon rank-sum test p=3.868e-05). Despite this trend, both the SMP and BDLP were heavily skewed towards infrequently observed phenotype associations, with more than half of all associations being observed ‘never’ or ‘rarely’ in both cases. P-values remain relevant with Bonferroni correction.

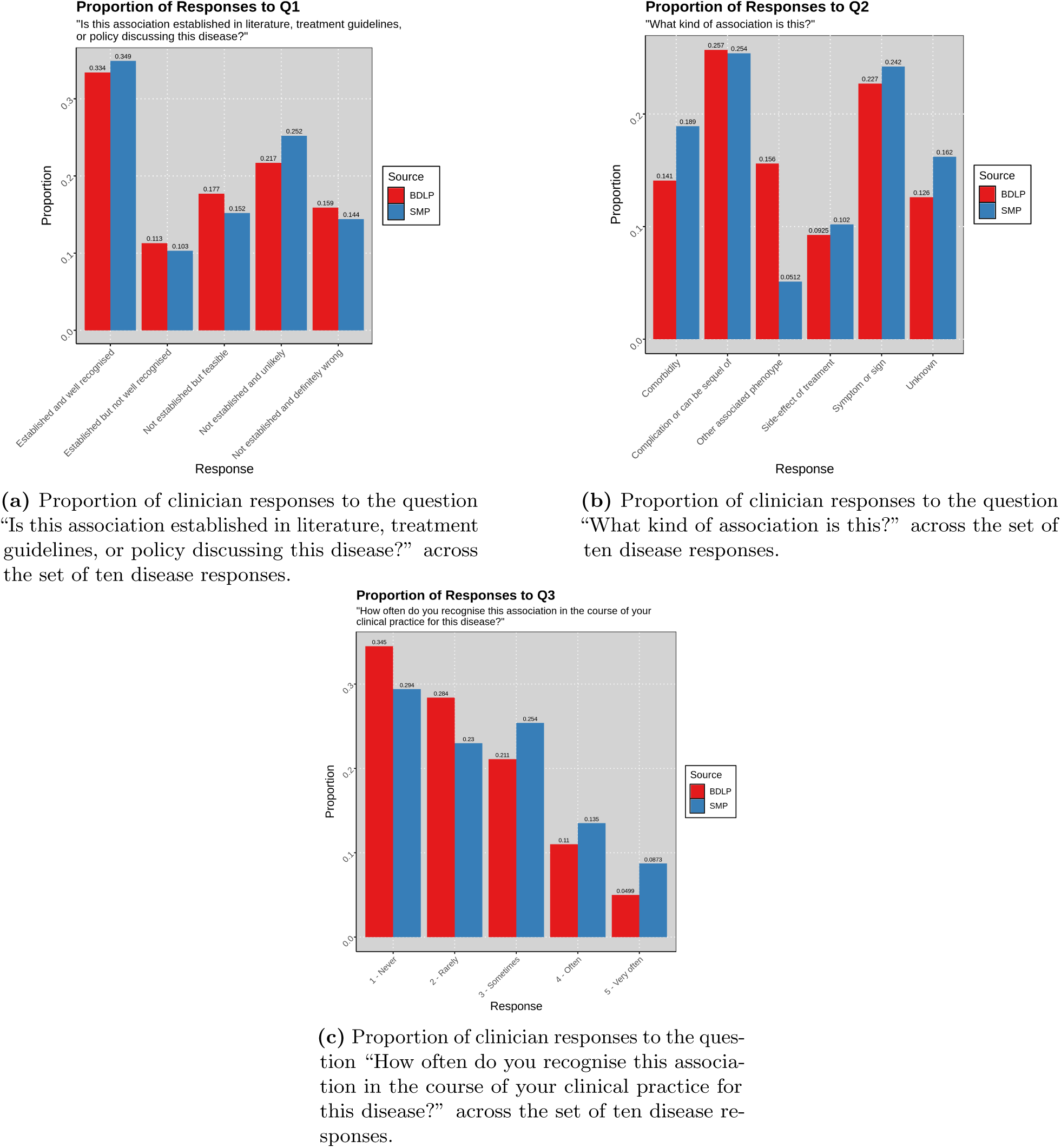

Using the results of the clinical review, we were also able to shortlist sets of phenotypes that are potentially novel. To do this, we isolated responses that were considered ‘Not established but feasible,’ of which there were 534, of which 93 were in the SMP and 465 in the BDLP, with the proportion of associations being similar (per Figure 4a) and 24 associations being recovered by both groups. We further minimised this set by removing the set of all associations that were recorded as being seen ‘never’ or ‘rarely.’ In the SMP, there were 23 phenotype associations meeting the criteria, and in the BDLP 79. The SMP contains a slightly higher proportion of this association subset than does the BDLP, with 0.034 and 0.03 respectively. We can also cross-reference this criteria for associations that were reviewed as not being well established, to provide a smaller set of hypothetical associations. These associations, of which there are 23, are shown in Table 4. The majority of 16 associations are concerned with fibromyalgia, with vasculitis, hypertrophic cardiomyopathy (HCM), and asthma also appearing. Of the 23, a majority of 18 were not present in the BDLP.

**Table 4.** The set of social media phenotype associations that were reviewed by a clinician as being ‘Not established but feasible’ but were seen at least ‘sometimes.’ These represent a small sample set of phenotypes that could potentially be novel, supported initially by the clinician account of their feasibility and prevalence. The “BDLP” column indicates whether this association was also found in the Biomedical Database and Literature Phenotype.

| Disease | Phenotype | BDLP | Seen In Clinic |
| --- | --- | --- | --- |
| Asthma (DOID:2841) | emphysema HP:0002097 | TRUE | 3 - Sometimes |
| HCM (DOID:11984) | respiratory insufficiency HP:0002093 | TRUE | 3 - Sometimes |
| HCM (DOID:11984) | abnormal blood ion concentration HP:0003111 | FALSE | 3 - Sometimes |
| HCM (DOID:11984) | abnormal vertebral morphology HP:0003468 | FALSE | 3 - Sometimes |
| Vasculitis (DOID:865) | abnormality of the foot HP:0001760 | FALSE | 3 - Sometimes |
| Vasculitis DOID:865 | diarrhea HP:0002014 | FALSE | 3 - Sometimes |
| Vasculitis (DOID:865) | abnormality of fluid regulation HP:0011032 | FALSE | 3 - Sometimes |
| Fibromyalgia (DOID:631) | urinary incontinence HP:0000020 | FALSE | 3 - Sometimes |
| Fibromyalgia (DOID:631) | tinnitus (HP:0000360) | FALSE | 3 - Sometimes |
| Fibromyalgia (DOID:631) | hearing abnormality HP:0000364 | FALSE | 3 - Sometimes |
| Fibromyalgia (DOID:631) | abnormal vertebral morphology HP:0000729 | FALSE | 3 - Sometimes |
| Fibromyalgia (DOID:631) | edema HP:0000969 | FALSE | 3 - Sometimes |
| Fibromyalgia (DOID:631) | bruising susceptibility HP:0000978 | FALSE | 3 - Sometimes |
| Fibromyalgia (DOID:631) | skin rash HP:0000988 | FALSE | 3 - Sometimes |
| Fibromyalgia (DOID:631) | gait disturbance HP:0001288 | FALSE | 3 - Sometimes |
| Fibromyalgia (DOID:631) | palpitations HP:0001962 | FALSE | 3 - Sometimes |
| Fibromyalgia (DOID:631) | nausea HP:0002018 | TRUE | 3 - Sometimes |
| Fibromyalgia (DOID:631) | vertigo HP:0002321 | TRUE | 3 - Sometimes |
| Fibromyalgia (DOID:631) | sciatica HP:0011868 | FALSE | 3 - Sometimes |
| Fibromyalgia (DOID:631) | night sweats HP:0030166 | FALSE | 3 - Sometimes |
| Fibromyalgia (DOID:631) | diminished ability to concentrate HP:0031987 | FALSE | 4 - Often |
| Fibromyalgia (DOID:631) | low levels of vitamin d HP:0100512 | TRUE | 3 - Sometimes |
| Fibromyalgia (DOID:631) | food allergy HP:0500093 | FALSE | 3 - Sometimes |

## Discussion

We described the creation of a social media derived phenotype model for a set of diseases, in the form of associations between those diseases and phenotypes. We also collected and combined an associative database from multiple literature analyses and experimental databases, representing the academic perspective on diseases. We linked a set of diseases from these two sets, comparing and contrasting them, with the hypothesis that they would be notably different, reflecting the differences in perspective between the academic community and the public. The results showed that while the social media phenotype recapitulated the academically-derived phenotype, the majority of social media derived associations (72.87%) did not appear in the biomedical database and literature derived phenotype model, despite a similar proportion of associations being found to be valid by clinical reviewers. We can, therefore, conclude that the phenotypes derived from social media represent strikingly different perspectives on disease.

### Clinical Perspectives

The clinical review of a subset of diseases showed that from the clinician’s perspective, the social media derived phenotypes and biomedical and literature derived phenotypes were similarly valid, or well established. They also showed that for those diseases, the social media derived phenotypes were seen significantly more often in the course of practice. That the clinical review results showed similar validity, as well as the substantiation of novel associations included subsequently through literature review, indicates that phenotypes novel to the SMP are not generally completely unmentioned in literature. Nevertheless, they did not appear together enough to be considered associated, and therefore did not appear in the relevant associative dataset. As such, these novel associations may constitute hypotheses or additional evidence for phenotypes that should be considered for further exploration.

In terms of the subset of phenotypes that were not known to clinicians, these can be interpreted in different ways. Where associations are novel to clinicians but appear in literature, it is likely that there is some evidence or association that has not found its way yet into the level of clinical translation that would mean that a clinician knows about them. This is perhaps why there are fewer ‘feasible’ relationships here. In the interests of exploring further the different knowledge and priorities encoded in datasets discussing healthcare entities, similar investigations could be undertaken on routine clinical datasets. These could involve textual analyses, but also analysis of supported by a common phenotype profile representational schema [30]. The necessity of exploring the clinical perspective is illuminated by the results of the clinical analysis. While they confirmed that the social media-derived phenotype was similarly aligned to, with a similar distribution of answers for across the evaluated diseases, they showed that neither BDLP or SMP were particularly well-aligned with clinical experience, with more than half of associations in both cases not being seen more than ‘rarely,’ and more than 30% in both cases being considered ‘definitely wrong’ or ‘unlikely.’

Using the results of the clinical review, we were able to illustrate a small set of potentially novel SMP phenotypes by identifying those that were found to be non-established, feasible, and seen in clinic at least sometimes. The dataset, however, includes a greater number of phenotypes of potential interest, for example those that are seen rarely from the clinician’s perspective may nevertheless be valid. In the subsequent section, we provide a narrative review of those phenotypes and their feasibility for two diseases.

#### Fibromyalgia

Fibromyalgia is characterised by chronic widespread pain alongside a number of non-pain features such as intrusive fatigue, poor refreshment from sleep, poor concentration and short term memory, and hypersensitivity to visual, auditory, and tactile stimuli [10]. Although the exact mechanisms remain unknown, a wealth of evidence shows that altered central nervous system processing can drive or maintain chronic pain in the absence of peripheral nerve or tissue damage [18]. It is possible that some of the sensory non-pain related symptoms highlighted in the current study, such as tinnitus, palpitations, vertigo, nausea are also due to central augmentation but have received less attention in the literature. For example, some studies have shown a mismatch between self-reporting of palpitations amongst patients with fibromyalgia, compared to healthy controls, in the absence of any significant differences in objective cardiac measurements [7, 20]. Autonomic dysfunction has also been proposed as a potential mechanism for some for the symptoms identified by the SMP including palpitations and skin manifestations, but objective data to support this theory is currently lacking [7, 27, 44]. Diminished concentration is a well-established phenotype of fibromyalgia, and was incorrectly marked as ‘not established’ during the validation phase.

Hearing and balance related symptoms were also identified by the SMP, whereas research in this area is also lacking. Preliminary data suggest a higher handicap relating to the presence of dizziness in patients with fibromyalgia compared to controls [49]. Furthermore, a recent, small-scale cross sectional study of the impact of tinnitus in fibromyalgia provides preliminary support for the link with central sensation as the severity of tinnitus was associated with severity of overall symptoms of fibromyalgia, and poorer quality of life [15]. Self-reported hearing loss, amongst other sensory symptoms, has also been previously shown to be more prevalent amongst patients with fibromyalgia compared to those with other rheumatic conditions, adjusting for age and sex [67]. Whilst this may represent central augmentation, increased reporting of non-sensory symptoms, including easy bruising, highlight the fact that other mechanisms must be involved.

Some of the unique phenotypes identified by the SMP may reflect the diversity in body systems effected by fibromyalgia, with some symptoms falling beyond the currently acknowledged causal mechanisms. For example, urinary incontinence has been linked to weakened pelvic floor muscles in patients with fibromyalgia, which is in turn related to the presence of lower urinary tract symptoms such as urinary incontinence [19]. Although the exact mechanisms are not known, early data suggest impairment of the nerve roots supplying the urinary and anal sphincters [28]. Similarly, a systematic review and meta-analysis has shown that people with fibromyalgia walk with a cycle of shorter length and lower frequency, producing a slower gait [13]. In addition, patients have a higher rate of perceived exertion with the 6 minute walking test [24]. Although abnormalities in individual vertebrae morphology has not been shown in the literature, abnormal spinal alignment has been reported [2, 36] and further investigation is warranted.

In contrast to many of the symptoms described above which generally lack attention in the medical literature, the potential relationship between vitamin D and fibromyalgia has been extensively studied [17, 68]. In theory the mechanisms by which vitamin D may be relevant in fibromyalgia include effects on skeletal muscle, neurotransmitters and neuronal regulation [35]. The issue is that observational and supplemental studies have produced conflicting results. This may be due, at least in part, to the heterogeneity of fibromyalgia and the difficulty in adequately capturing a meaningful change in symptoms at the individual level.

Although there has been much interest in the effect of diet on fibromyalgia, the current evidence base remains inconclusive. A survey of 101 patients with fibromyalgia suggested that the self-reported frequency of food allergy is likely to be higher than the general population [6]. Whilst further evaluation is needed, this is consistent with the observation that patients are more likely to report allergies more generally, and may also represent hypersensitivity rather than true allergy similar to drug hypersensitivity that is seen in fibromyalgia [34]. It also suggests that fibromyalgia patients are seeking out dietary measures which is understandable given the lack of pharmacological treatments and focus on lifestyle measures more generally.

#### Hypertrophic cardiomyopathy

Although it is difficult to envisage electrolyte abnormalities (synonymous with blood ion concentration abnormality) occurring in unmedicated patients as a direct result of hypertrophic cardiomyopathy, it would be feasible for patients treated for heart failure or in those with concomitant renal disease. The relationship between HCM and electrolyte abnormalities may be overlooked in the scientific literature due to the focus on cardiac-specific biomarkers, such as B-Type natriuretic peptide (BNP) and Troponin. Electrolyte abnormalities are related to prognosis in patients with heart failure from any cause [5]. The idea that routine blood tests, which international guidelines recommend are taken at a patient’s initial assessment [4], could help refine the understanding of disease trajectory opens an immediate avenue for investigation.

Meanwhile, the appearance of respiratory insufficiency as unestablished appears to be a labelling error, as breathlessness due to heart failure is well-described in the scientific literature. Researchers describe most cohorts according to their levels of breathlessness using the New York Heart Association (NYHA) classification.

Abnormal vertebral morphology may be connected to hypertrophic cardiomyopathy through a neuromuscular condition called Freidrich’s Ataxia, where patients are affected by scoliosis and an HCM-like cardiac phenotype [4]. The relationship may be describing affected individuals who have not yet received a formal diagnosis of Freidrich’s Ataxia. A link to HCM itself would be surprising and challenging to explain since changes in this condition are caused by abnormalities of the sarcomere and are confined to the heart.

### Thematic Analysis and Public Contexts

Using the ontology to contrast the themes and categories of the associations, it was clear that the social media phenotype was heavily skewed towards constitutional symptom phenotypes. Constitutional symptoms are defined in the Human Phenotype Ontology as “*[…] indicating a systemic or general effect of a disease and that may affect the general well-being or status of an individual*”, with further guidance on the classification specifying that the category is defined by phenotypes that affect patient quality of life. The largest contributor to these new associations was, by far, pain, and its more specific subclasses, which was also the largest contributer to overall novel associations, at 6%. Other large contributions from constitutional symptoms came from phenotypes including fatigue, impairment of activities of daily living, night sweats, and indigestion. Upon further investigation of constitutional phenotypes and their accordance to disease areas, we found that there was a greater focus on abdominal, endocrine, and reproductive system disease.

Despite the large number of additional pain-related associations, we showed that most of these were concentrated around more general, less specific, phenotypes. Conversely, the BDLP pain phenotypes were more distributed across the full set of pain phenotypes defined in the Human Phenotype Ontology. We suggest that this is partially resulting from the public not knowing more advanced and technical medical terms for more specific kinds of pain, but also that these more specific terms are not necessarily relevant to the context in which a symptom is being discussed on social media. For example, there were no associations for ‘precordial pain,’ but there were mentions of its more general parent, ‘chest pain.’ In this case, ‘precordial’ is a technical term the many members of the public may not be familiar with, and the use of it in a social media conversation does not necessarily communicate an informative difference, at the cost of wider interpretability. Conversely, the specifying difference between chest and precordial pain may be highly relevant in the context of an academic study or clinical care.

Some relatively simple terms were also not represented in the SMP, however, such as ‘wrist pain,’ and we believe that this points towards our methodology. An area of difficulty in comparing these datasets is our use of a relatively stringent significance requirement for consideration of an association in the analysis. This is closer to the Pilehvar et al methodology, which used a false discovery cut-off with a Fisher exact test. Meanwhile, the Kafkas et al work provided all associations with positive NPMI scores, reporting that a threshold of 76 phenotypes results in maximal similarity to manually curated associations. Our investigation used a very exclusive false discovery rate of 0.0005 in an effort to yield higher quality associations for our subsequent analysis, with the aim of discovering novel phenotype associations with high plausibility. Moreover, we did not want to align our significance testing with expert ground truth or recapitulation of biomedical databases, as the other two studies did, since our goal was to identify associations that are not necessarily aligned with this perspective on disease. Nevertheless, our approach yielded a similar distribution of validity between per our expert clinical analysis. In using this approach, however, we necessarily exclude many potentially valid associations, and this becomes more likely where concepts become more specific in the knowledge graph, which likely contributes to the concentration of SMP phenotypes on more general phenotypes (seen, for example, in Figure 3). While this makes it difficult to draw conclusions from differences in the appearance of more specific phenotypes between the two datasets, especially where they do not appear in the more strict SMP, it does not preclude the thematic analyses upon which we have focused in this paper, and places a greater interest on associations that were identified. A wider exploration of methods for scoring co-occurrence should be considered as future work, as well as methods for identifying and evaluating interesting associations, even where they fall below a relatively high significance threshold. Ultimately, multi-contextual phenotype models should be developed using equivalent methodologies for a more fair comparison, though we believe the current study provides initial evidence that a social media phenotype model is a valuable resource, worthy of further investigation.

The White Swan data were sourced from a wide range of social media sources. These included general social media such as Twitter, but also Reddit, which is organised into many topic-based sub-fora, as well as other sources. As such, it is highly likely that there are many sub-contexts expressed in these data, for example differences between discussions of disease informed more by a more general social opinions, and those informed by the more specific and technical understandings exhibited by those with direct experience of a disease. For example, previous work found that while patients used different language and had different priorities, they knew and used advanced medical terminology in online conversations [50]. Further exploration should be undertaken to explicate these contexts, with differences being drawn between the understandings and perspectives held in different social spheres of the web.

There were entire facets for which the BDLP is overall more connected than the SMP. For example, it describes a far greater number thoracic cavity phenotype associations. We suspect that this could relate to the number of layperson synonyms defined in the HPO for those phenotypes, since those synonyms contributed to the text mining vocabulary used by this study. A study describing development of layperson synonyms in HPO reported 0% coverage for the thoracic category [63]. To a lesser extent, voice phenotypes are also under-represented in the SMP, despite that group being reported as having 44% layperson synonym coverage. This perhaps speaks to the relatively small size of the voice facet of HPO, which is largely concerned with highly technical terms, whose layperson synonyms form complicated compound phrases which are unlikely to be found in conversational text, e.g. ‘weakness of the vocal cords.’ Other components, such as ‘cries,’ are mostly associated with babies, who are unlikely to be expressing themselves on social media. Facets that are under-expressed in the SMP could represent those patients are less aware of, or less interested in, or they could indicate poorer alignment of the vocabulary with the language they use.

At a more basic level, the inherently error-prone nature of text-mining and large-scale association mining, as well as the shift in language meaning across contexts, mean that extracted disease-phenotype associations may not actually reflect true biomedical relationships. For example, the phenotype anorexia (HP:0002039) is defined as “A lack or loss of appetite for food (as a medical condition),” which is distinct from the disease anorexia nervosa (DOID:8689). This distinction may be lost in a public context, where ‘anorexia’ is often used as a referent for the disease, and more rarely for the phenotype of poor appetite. These limitations are, however, a component of any co-occurrence approach to determining relationships between biomedical entities, with scientific literature also referring to the disease with the unqualified ‘anorexia’ in some cases [12]. Further complicating this example is ‘anorexia’ being a substring of ‘anorexia nervosa,’ meaning that in many text mining approaches, all instances of ‘anorexia nervosa’ in text would also be labeled as an instance of anorexia. Improvements to the text mining methodology could also mitigate issues with limitations to the use of formal terminologies for text mining. Particularly, the transactions provided by White Swan were determined using a rule-based NLP system, and therefore required exact mentions of labels included in the vocabulary to link an entity. This approach was shared by the Kafkas et al approach. State-of-the-art approaches to text mining in a healthcare context often employ contextual embedding similarity to identify and link mentions using labels not explicitly defined in the underlying vocabulary [41], and the Pilehvar et al approach used such a method. The employment of this kind of method would aid in linking mentions that are not pre-defined in the relevant vocabularies, which would be especially beneficial in the use-case of picking up mentions from social media, although these approaches come at the cost of an increased error surface for erroneous annotations, and additional complications in determining appropriate cut-offs. In the context of this experiment, we were not able to modify the text mining methodology, since results were provided to the authors by White Swan, using their internal text mining framework. In a similar manner to the more strict statistical boundary used in our approach, a rule-based approach to NER makes it difficult to infer from the absence of phenotypes from the SMP, but does not affect interpretation of their appearance, especially where those associations do not appear in the literature dataset.

For these reasons, ultimately, while our work identifies a large number of hypothetical relationships between diseases and phenotypes that are not reflected in current academic databases, further work must be done to explore them and to identify what, if any, scientific or clinical utility they have. This limitation is also relevant to the associations recovered by the other studies we explored that make up the BDLP, and we anticipate a programme of research that surrounds the alignment, comparison, and evaluation of multi-contextual disease phenotypes in a single methodological context.

One previous study has also identified a critical need for correlating digital phenotyping data with epidemiological data [47]. Recent efforts such as BioLink [62] aim to formalise and harmonise biomedical entity associations, however they do not include extensive vocabularies for text mining, and do not include a rich metadata language for describing the derivation and provenance of calculated associations. In our review, while clinicians were largely able to categorise all phenotype associations into a small number of categories, with a relatively small number percentage of associations being marked as ‘other’ or ‘unknown,’ this required a lot of manual work, and the automated inference of the nature of these relationships from text could be considered a task for future work.

We also envision that these hypothetical relationships can be used as prompts for patient interaction and involvement, building an integrated evidence base for introducing changes to clinical practice that more closely reflect and serve public and patient priorities. These processes can also be used to query the exact nature of the relationships and perspectives being explored, ensuring that they are more fully understood. We anticipate that the use of deep phenotyping data from a range of multi-contextual resources can be employed as a contributing device in an increasing drive towards patient-centred research and care.

## Conclusions

We developed a social media derived phenotype model of disease to represent public and patient perspectives on disease and their signs and symptoms. We have demonstrated that this phenotype model expresses a significantly different perspective than that expressed by biomedical databases and literature. Moreover, we identified a large number of novel associations that were not represented in the biomedical and literature model. We anticipate that this knowledge resource can contribute to an improved understanding of the human diseasome across healthcare research and implementation, and that analysis of diverse data sources can contribute to a fairer and increasingly patient-centred approach to medicine.

## Methods

The methods used to produce our results are available online at https://github.com/reality/sd_paper.

### Dataset

We obtained a database of transaction data describing mentions of diseases and phenotypes from White Swan, a charity based in the United Kingdom. White Swan queried text data from a range of sources including social media and patient voice fora for posts describing a set of diseases of interest. Those posts were internally analysed for mentions of Human Phenotype Ontology (HPO) and Disease Ontology (DO) classes, using vocabulary identified for those phenotypes and diseases across identifiers across biomedical ontologies, using the method described in Slater et al [55]. The dataset covers a period of 1st November 2019 to 1st November 2021.

### Social Media Phenotype (SMP)

We propagated mentions of classes for phenotypes across the set of transaction records to superclasses. For example, any mentions of ‘low back pain’ would also be considered mentions of ‘back pain,’ and ‘pain.’ As described by Kafkas et al, this was achieved using a modified measure of Normalised Pointwise Mututal Information (NPMI) that takes into account subsumptive hierarchy, shown in Equation 1. Subsumption was identified using the ELK ontology reasoner [37]. We did not propagate disease mentions, due to the limited number of diseases being examined causing potential bias to arise due to incomplete composition from subclasses.

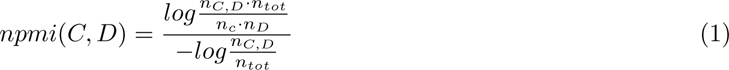

We calculated NPMI values for every combination of disease and phenotype. We then used a Monte Carlo bootstrapping approach to simulate a randomised transaction set with the same size and shape 2,000 times, using NPMI values calculated from those simulations to identify p-values for every association. We confirmed normality of scores across the simulations visually.

We excluded from further consideration any associations with a non-positive NPMI value, since this study is concerned with positive associations. We also excluded diseases and phenotypes that appeared in fewer than 0.01% of transactions. We did this to avoid skewed NPMI values for associations involving classes that appeared very infrequently. The remaining set of associations comprises the SMP. We then calculated values [60] for all SMP associations, using an acceptable false discovery rate of 0.0005, with associations meeting this threshold being considered significant in the context of this study. We also performed a review of phenotypes included in associations to identify any with associated labels that would be unlikely in a public context to imply a similar phenomenon to the actual phenotype (e.g. plethora (HP:0001050)), or that had erroneously associated labels (no social interaction (HP:0008763) also had label ‘social interaction’). These phenotypes were discluded from the final set of associations and therefore removed from further consideration in our experiment.

### Biomedical Database and Literature Phenotype (BDLP)

To construct a resource representative of existing background knowledge resources describing biomedical databases and literature, we collected phenotypes from several resources: the set of text-mined literature associations from Kafkas et al, semi-automatic disease-phenotype associations from Kafkas et al, and the text-mined literature derived phenotypes from Pilehvar et al [33, 51]. All of these datasets use HPO to describe phenotypes. However, Kafkas et al used ICD-10 for diseases, while Pilehvar et al used MONDO. We linked diseases to DO using the database cross references defined in MONDO and DO. For unlinked diseases we developed manual mappings, where possible, and contributed these associations back to DO. We consolidated phenotype associations from the three sources listed above for all diseases with a cross-reference to DO. These consolidated associations made up the Biomedical Database and Literature Phenotype (BDLP) dataset.

### Identifying Novel Associations

So as to find a laconic intersection of these associations, we determined a subset of social media derived associations for each disease. We labeled all social media disease-phenotype associations with any more specific association in either the BDLP or the SMP for that disease, or an equivalent association in the BDLP for that disease. In this way, we identify a subset of social media phenotypes for each disease that are significant, maximally specific, and distinct from those in the literature derived set. We developed a website at http://phenotype.digital/ for exploring and comparing the phenotypes of diseases across the two contexts. ChatGPT was used to aid in the development of the website front-end, which was implemented in NodeJS.

### Clinical Review

We sought clinical collaborators to review associations. All reviewers are consultant clinicians with an active practice in the disease they reviewed in the UK. Reviewers were blinded to the source of the associations, and their order was randomised. The questions asked were:

1. Is this association established in literature, treatment guidelines, or policy discussing this disease? (established and well recognised, established but not well recognised, not established but feasible, not established and unlikely, not established and definitely wrong).
2. What kind of association is this? (symptom or sign, comorbidity, complication or can be sequel of, side-effect of treatment, other associated phenotype, unknown)
3. How often do you recognise this association in the course of your clinical practice for this disease? (1 - never, 2 - rarely, 3 - sometimes, 4 - often, 5 - very often)

The reviews were conducted via a web application implemented into the digital phenotype website, and which was developed to permit additional review and collection of expertise in the future. The diseases reviewed were bronchiectasis (DOID:9563), pancreatic carcinoma (DOID:4905), systemic lupus erythematosus (DOID:9074), skin carcinoma (DOID:3451), dermatitis (DOID:2723), asthma (DOID:2841), hypertrophic cardiomyopathy (DOID:11984), pulmonary embolism (DOID:9477), vasculitis (DOID:865), congestive heart failure (DOID:6000), chronic obstructive pulmonary disease (DOID:3083), and fibromyalgia (DOID:631).

### Evaluation and Analysis

To measure how well the SMP semantically recapitulated the BDLP, we calculated an Area Under the Curve (AUC) by ranking semantic similarity scores calculated using the Resnik method [53], comparing the phenotype profiles of each pairwise combination of matched diseases in the BDLP and SMP. Semantic similarity was calculated using the Semantic Measures Library [23]. We used the Klarigi tool to perform compositional analysis of phenotypes [57], automatically correcting inclusion scores for subsumptive phenotype relationships implied by the underlying ontology. A combination of Groovy and R were used to perform evaluation. Figures were produced using the ggplot library.

## Funding

The authors acknowledge support from the NIHR Birmingham ECMC, NIHR Birmingham SRMRC, Nanocommons H2020-EU (731032), NIHR BBRC, MRC HDR UK (HDRUK/CFC/01), KAUST OSR (URF/1/3790-01-01), and MRC (MR/S003991/1), MAESTRIA (Grant agreement ID 965286), HYPERMARKER (Grant agreement ID 101095480), PARC (Grant Agreement No 101057014) and the MRC Heath Data Research UK (HDRUK/CFC/01) and HDRUK midlands regional community project [QQ2], initiatives funded by UK Research and Innovation, Department of Health and Social Care (England) and the devolved administrations, and leading medical research charities. The views expressed in this publication are those of the authors and not necessarily those of the NHS, the National Institute for Health Research, the Medical Research Council or the Department of Health. PNS acknowledges the support of The Alan Turing Institute.

## Acknowledgements

We would like to thank White Swan for their provision of data and help with analysis, with particular thanks to Beth Fordham. We thank Dr. Jackie Williams for discussions and suggestions surrounding methodology and impact, and Lauren Cooper for help in project management and recruiting clinical collaborators. In addition to the named authors, we are extremely grateful to the additional review respondents: Dr. Keith Roberts, Prof. Alice Turner, Dr. Benjamin Mulhearn, Dr. Jess Ellis, Dr. Anita Sullivan, Dr. Clara Green, Dr. Caitlin Stevens, Dr Ser-Ling Chua, Dr Agustin Martin-Clavijo, and Dr. Asgher Champsi.

## Data availability

The result data and intermediate data are available via https://github.com/reality/sd_paper and https://phenotype.digital/.

## Code availability

The code for experiments described in this paper is available from https://github.com/reality/sd_paper.

## Ethics approval and consent to participate

This project was reviewed by the ethics committee at the University of Birmingham, and granted full ethical approval with identifier ERN 2022-0241 and ERN 0241-Jun2023 amendment.

## Competing interests

James Wright is an employee of White Swan, who provided the dataset for this study. Otherwise, the authors declare that they have no competing interests.

## Consent for publication

Not applicable.

## Author contributions

**KS** conceived of the study, designed and implemented the experiment, performed the data analysis, and drafted the manuscript. **PNS** contributed to experimental design and clinical interpretation of results. **JW** performed data pre-processing and contributed to subsequent data analysis. **AI** contributed to clinical interpretation of results. **WB** contributed to clinical interpretation of results, and development of questionnaire design. **FA** contributed to the evaluation of results and study presentation. **GVG, PC** supervised the study, contributed to experimental design, and contributed to the manuscript. All authors approved the manuscript for submission.

## Notes

### Author Declarations

Science, Technology, Engineering and Mathematics Committee Ethics board of University of Birmingham gave ethical approval for this work with ERN_2022-0241 and amendment ERN_0241-Jun2023.

